# A micro-costing of latent tuberculosis infection testing and treatment in adults in secondary care in England

**DOI:** 10.1101/2025.01.29.25321354

**Authors:** Laura Snoad, Juan Vesga, Marc Lipman, Martin Dedicoat, Rishi K Gupta, Ibrahim Abubakar, Susan Dart, Thomas Gorsuch, Heinke Kunst, Sarah Mungall, Ryan Noonan, Peter J White

## Abstract

**Background:** Latent TB infection (LTBI) screening and treatment of population groups at high risk for TB is part of England’s Tuberculosis Action Plan, which has succeeded in increasing testing volumes. However, analysis of the cost-effectiveness is hampered by a lack of detailed information on health service costs.

**Methods:** We surveyed clinics with large volumes of activity, located in the London, West Midlands, South East & South West, and North West England TB Control Board areas. These represent three-quarters of England’s TB diagnoses. We mapped clinical pathways and determined costs of staff time, testing, and drugs.

**Results:** The clinics use interferon gamma release assays (IGRAs) (Quantiferon-TB Gold Plus, T-SPOT.TB, or both) for LTBI testing, and the most common outcomes of the testing pathways are asymptomatic patients testing negative (costing £64.11 per patient, range: £49.08-£81.02) or positive (costing £142.86(£120.95-£174.68)). Symptomatic patients, testing positive or negative, have higher costs (£124.35(£101.29-£138.23)). The estimated cost per patient diagnosed with LTBI is £458.32(£362.49-£571.47). Treatment costs depend on the regimen and the amount of monitoring the patient requires, e.g. 3 months daily Isoniazid and Rifampicin costs £192.68(£140.43-£255.32) for most patients, and £238.62(£157.47-£346.03) for those requiring more monitoring. 6 months daily Isoniazid costs £411.04(£362.56-£474.25). 4 months daily Rifampicin costs £226.97(£190.15-£270.73). One clinic uses 3 months of weekly Isoniazid and Rifapentine for some patients, costing £559.05.

**Discussion:** These are the first detailed cost estimates of latent TB screening and treatment, and will enable improved assessment of cost-effectiveness, and allocation of resources.

## INTRODUCTION

Screening and treatment for Latent Tuberculosis Infection (LTBI) is part of both the World Health Organization’s End TB Strategy^1^ and England’s Tuberculosis (TB) Action Plan^2^. However, a large majority of people identified as having LTBI do not progress to active TB disease, even without treatment. With limited resources for public health, it is vital that LTBI interventions are cost-effective. At present, assessment of cost-effectiveness is hampered by a lack of data regarding the costs of current procedures. Accurate cost information is essential for ensuring value for money of LTBI interventions and that the correct amount of funding is allocated to screening and treatment. Although clinical guidance sets out testing and treatment protocols^3^, how that is delivered varies for different categories of patients and for different NHS trusts, with different cost implications.

This study estimated costs of LTBI testing and treatment in secondary care, focusing on adult new entrants from high-TB-burden countries (defined by the NHS as countries with a TB incidence ≥150/100,000 per year and countries from Sub-Saharan Africa^4^) and adult contacts of infectious cases, the population groups which represent the large majority of those tested and treated for LTBI. The study examined clinics in England, mapping their LTBI care pathways to estimate costs of staff time, tests and drugs.

## METHODS

Using data from UKHSA’s TB Surveillance system^5^, we identified the four TB Control Board areas in England (Figure 1) with the most TB diagnoses, and then approached clinics in these areas to participate. In this report the clinics are anonymised.

**Figure 1:**
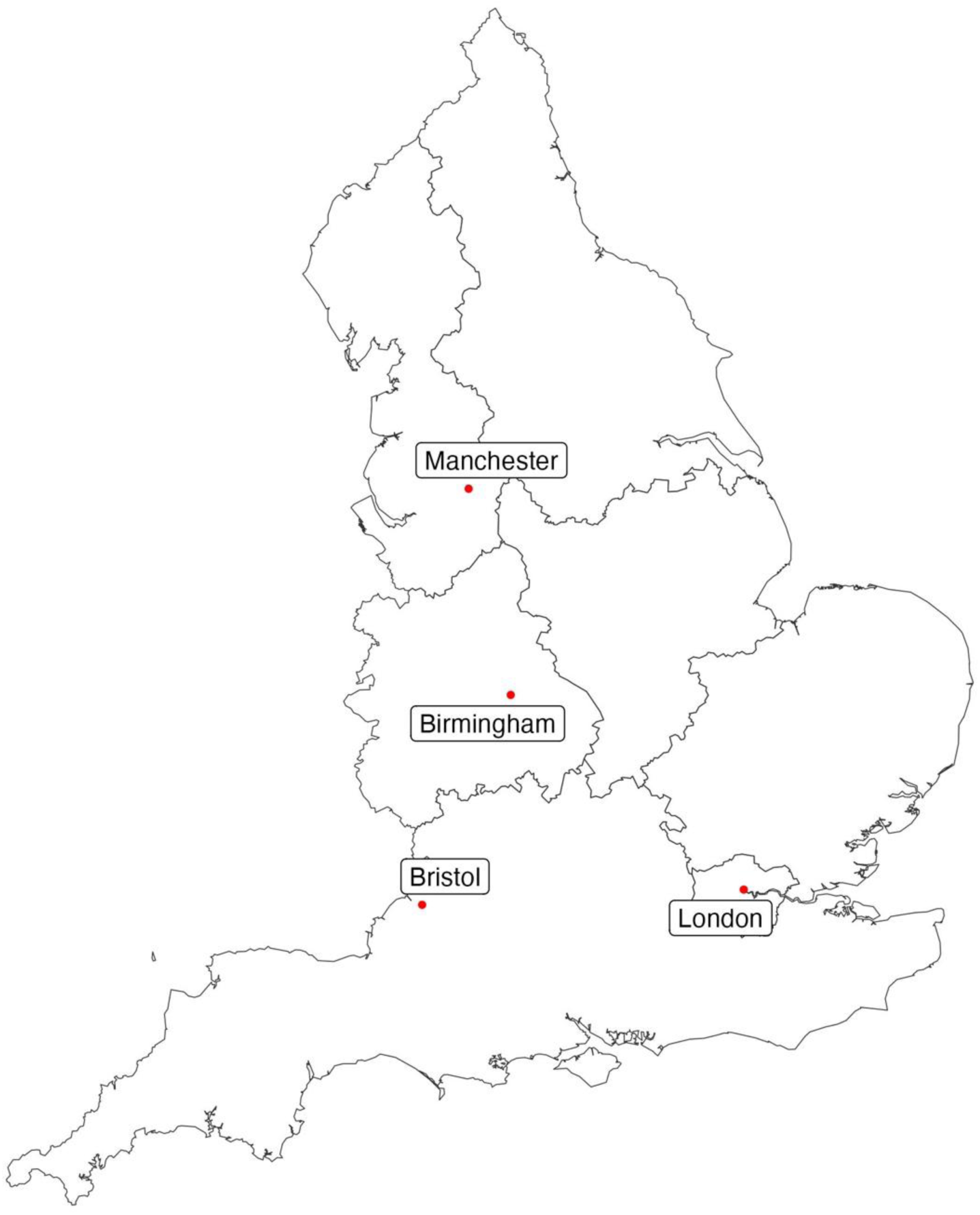
Map showing the seven TB Control Boards in England and the locations of the five clinics in the study.

We defined testing pathways as beginning with initial patient contact and ending with the patient having been determined either to have LTBI (and be invited to begin treatment) or not to have LTBI. The small proportion of patients who were ultimately diagnosed with active TB were not considered in this analysis. We defined the treatment pathway as beginning when a patient diagnosed with LTBI attends a consultation to discuss starting treatment, and ending with the completion of the drug regimen.

For each clinic, there was an initial interview with the lead clinician to define that clinic’s LTBI testing and treatment pathways. This identified the types of patients screened and treated by each clinic, how (if at all) the pathway differed for different types of patient, which LTBI diagnostic tests are used (and if more than one then under what circumstances each is used), the LTBI drug regimens used, and how treatment monitoring is conducted. Pathway diagrams for each clinic were then created. These were used as a visual aid at a second interview with the lead clinician or other key informant to determine the grade(s) of staff and the amount(s) of their time required for each stage of the pathway. Uses of staff time included attending to the patient, reviewing results, and administrative tasks such as communicating results and booking appointments. The tests performed (and drugs dispensed, where relevant) at each stage were also recorded. In some cases, the grade of staff who performs a particular task varies according to availability; here we used the average cost of the relevant grades assuming equal probability. In other situations, where the grade of staff depends on patient characteristics (e.g. potentially having active TB in the screening pathway or being thought to be at higher risk of drug-induced liver injury (DILI) in the treatment pathway), these were treated as distinct pathways for cost calculations.

Per-minute costs for different grades of staff were calculated by dividing annual costs of employment by the number of minutes worked per year. Annual costs of employment were estimated from published pay scales, using the mid-point of each salary range for UK (excluding London), Outer London, and Inner London^6,7,8,9,10,11^. Additional costs of employer pension contributions^12^ and employer National Insurance contributions^13^, the Apprenticeship Levy^14^, and indirect costs^15^ were included. The details of the calculation are in the Appendix.

Most clinics test for LTBI using an interferon gamma release assay (IGRA) either Quantiferon-TB Gold Plus (QIAGEN) or T-SPOT.TB (Oxford Immuntoec, now trading as Revvity) exclusively, but where both tests are used in the same clinic, clinicians were asked to estimate what proportion of the time they used each test, so that a weighted average cost per test could be calculated.

Costs for each treatment regimen (for a 70kg adult) were based on the British National Formulary^16^ or eMIT database^17^, selecting the cheapest option from either. Unit costs of tests were available from Clinic 1. Detailed values for costs can be found in the Appendix.

Costs are reported in 2024-25 GBP £.

## RESULTS

Five clinics participated, from the London, South East & South West, West Midlands and North West TB Control Boards, which represent three-quarters of TB diagnoses in England. None of these clinics use TST to diagnose LTBI, except in children or in rare cases where the initial IGRA result was thought to be unreliable, and so TST was not included in this analysis.

Figure 2 illustrates typical routes through LTBI testing and treatment services (pathways for each clinic are in the Appendix). Patients travel through the care pathway based on the results of an initial symptom screen. Asymptomatic patients start their journey with an Interferon Gamma Release Assay (IGRA) test. Patients with a negative IGRA are usually told via phone call, while those with a positive IGRA are invited back to clinic for a chest X-ray and baseline tests, which may include liver and renal function tests, testing for HIV, Hepatitis B and Hepatitis C, Full Blood Count, C Reactive Protein, Bone Profile and Vitamin D tests (hence these tests being included in the “testing” pathway even though they pertain to treatment). Both outcomes are important to capture as communicating results to IGRA negative patients via phone requires staff time. IGRA positive patients with an abnormal chest X-ray or signs of extrapulmonary TB will start active TB investigation and so leave the pathway that we consider. For patients with a normal chest X-ray the next step is to discuss LTBI treatment. Appropriate treatment is also prescribed at this point.

**Figure 2:**
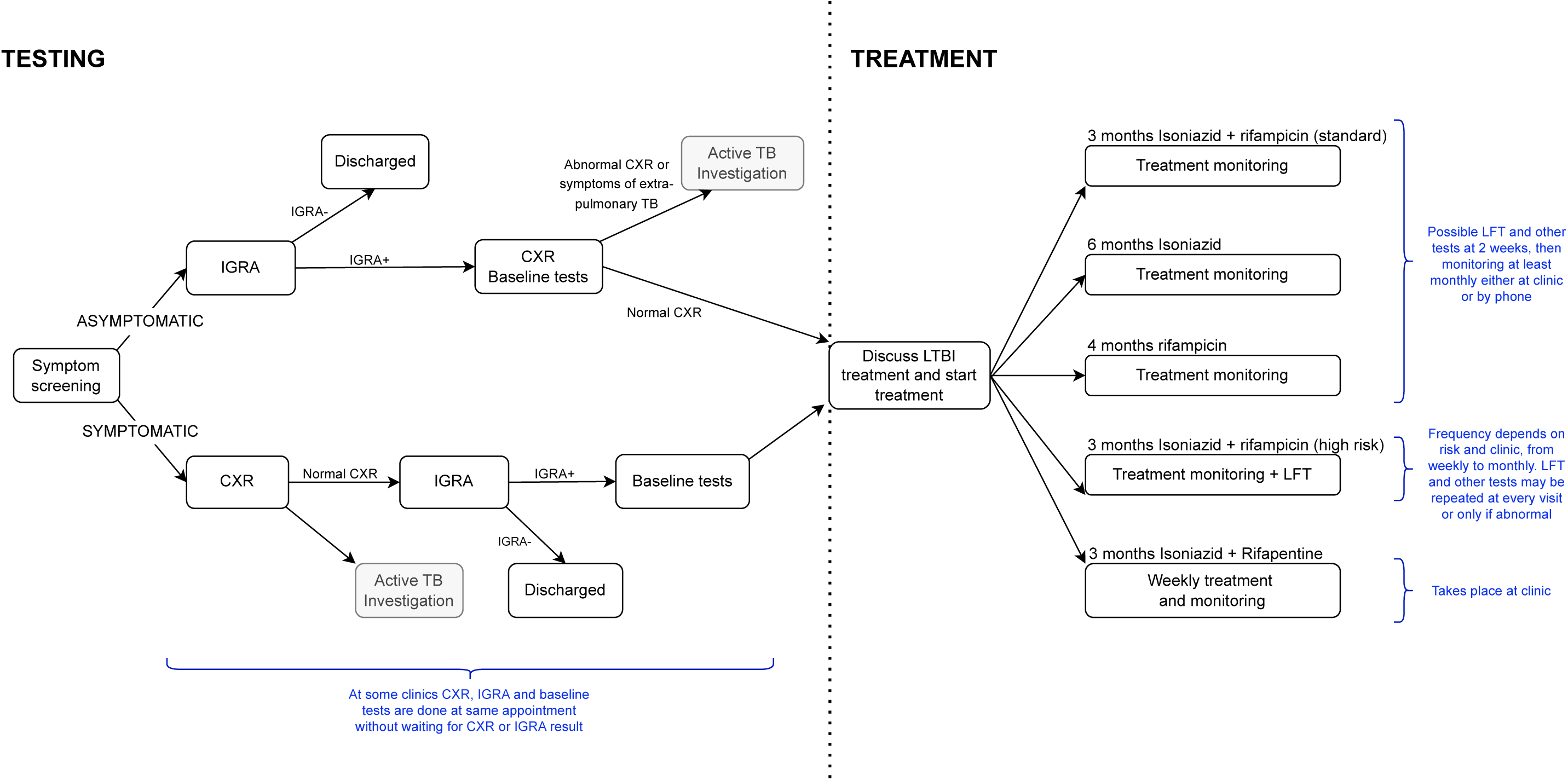
Schematic showing typical routes through LTBI testing and treatment services in NHS England Trusts where IGRA (Interferon Gamma Release Assay) tests are used. CXR: chest X-ray; LFT: liver function test.

Alternatively, for patients with symptoms consistent with active TB, the first step is to perform a chest X-ray. Some clinics wait for a normal chest X-ray result before conducting an IGRA test, but often in practice an IGRA test is conducted at the same time as the chest X-ray, alongside any baseline tests to exclude active TB such as C-reactive protein, to save staff time and reduce the risk of the patient’s not returning. Patients with a normal X-ray and positive IGRA then return to clinic to discuss their LTBI treatment and receive their prescription, in the same way as those patients who travelled down the asymptomatic pathway.

As standard a three-month course of Isoniazid and Rifampicin (3HR) is prescribed to treat LTBI^3^. Those at risk of drug interactions may be prescribed six months of Isoniazid monotreatment (6H), while four months of Rifampicin (4R) may be prescribed when clinicians wish to avoid isoniazid (e.g. in the context of peripheral neuropathy). Patients where adherence is a concern may be prescribed a three-month regimen of weekly isoniazid and rifapentine (3HP) taken at clinic, but this is rare; only one clinic in our study has access to Rifapentine and its availability in the UK is likely to remain limited. The frequency and nature of treatment monitoring depends on the regimen prescribed but also varies widely between clinics. Some clinics conduct a second liver function test at two weeks, and then further monitoring is done at regular intervals either in clinic or by phone. Patients deemed to be high risk because of abnormal baseline tests may return to clinic more frequently for monitoring.

### Costs of LTBI testing

One clinic was not included in the analysis of LTBI testing costs because its pathway combines primary and secondary care and so is out of scope of this analysis.

In the remaining four clinics, there are different pathways for asymptomatic patients undergoing LTBI screening, and for symptomatic patients who are assessed initially for active TB (Table 1). The outcomes of these pathways are (i) “Asymptomatic IGRA-” (asymptomatic patients who test negative by IGRA), (ii) “Asymptomatic IGRA+” (asymptomatic patients who test positive by IGRA and have LTBI diagnosed), (iii) “Symptomatic IGRA-” (patients with symptoms compatible with active TB but who have a normal chest X-ray and test negative by IGRA), and (iv) “Symptomatic IGRA+” (patients with symptoms compatible with active TB but who have a normal chest X-ray and test positive by IGRA so have LTBI diagnosed). Clinic 1 was able to provide the proportions of patients in these four categories in 2023, which were (i) 82%, (ii) 16%, (iii) 1%, (iv) 1%.

The most common testing outcomes, “Asymptomatic IGRA-” and “Asymptomatic IGRA+”, respectively cost a mean of £64.11 per patient using UK salary scales (range: £49.08-£81.02), and £142.86(£120.95-£174.68), with the large difference being due to the chest X-ray and baseline tests given to patients on testing IGRA-positive. Baseline tests may include liver and renal function tests, testing for HIV, Hepatitis B and Hepatitis C, Full Blood Count, C Reactive Protein, Bone Profile and Vitamin D tests, with the results informing the regimen prescribed and monitoring frequency. Patients presenting with symptoms of TB are much less frequent and have similar costs to “Asymptomatic IGRA+” patients, as they also have a chest X-ray and baseline tests. However, as these patients often have all these tests at one appointment (rather than at a follow-up appointment like “Asymptomatic IGRA+” patients), there is a lower staff cost of £124.35 per patient (range: £101.29-£138.23). All clinics reported the same costs for “Symptomatic IGRA-” and “Symptomatic IGRA+”. Using the proportions of patients in the four categories reported by Clinic 1, the estimated cost per patient testing positive (i.e. diagnosed with LTBI) is £458.32(£362.49-£571.47), which is the average cost of testing all patients divided by the proportion of patients who test positive.

Variation in cost for LTBI testing is largely driven by staff time. Clinic 3 consistently reported longer appointments, for example recording 30 minutes of radiographer’s time to conduct a chest X-ray, compared to 15 minutes at Clinic 2, 10 minutes at Clinic 1, and 5 minutes at Clinic 4. There is a slight variation between clinics in baseline tests done, with some clinics using vitamin D, bone profile and C-reactive protein tests. Clinic 1 uses Quantiferon 80% of the time and T-SPOT.TB 20%, Clinics 2 & 4 use only Quantiferon, and Clinic 3 uses only T-SPOT.TB.

### Costs of LTBI Treatment

All five clinics were included in the analysis of LTBI treatment costs. All clinics sampled provide the cheapest regimen (3HR) as standard, with other regimens prescribed as appropriate based on patients’ social factors, comorbidities, and risk of drug interactions.

Figure 3 shows the timeline for LTBI treatment for all regimens and clinics, while Table 2 shows costs of staff time, tests, and drugs. All five clinics reported prescribing three months of daily Isoniazid and Rifampicin (3HR) and six months of daily Isoniazid monotreatment (6H). Clinics 1,3 & 5 reported prescribing four months of daily Rifampicin (4R). Only Clinic 1 prescribes a three-month regimen of weekly isoniazid and rifapentine (3HP). For patients deemed to be at elevated risk of DILI^3^, all clinics reported a revised monitoring schedule. At Clinic 2, the standard treatment pathway is nurse-led clinic, but for patients with abnormal liver function test (LFT) results or who were symptomatic prior to LTBI diagnosis, treatment is doctor-led and therefore has higher costs (see Appendix for diagram).

**Figure 3:**
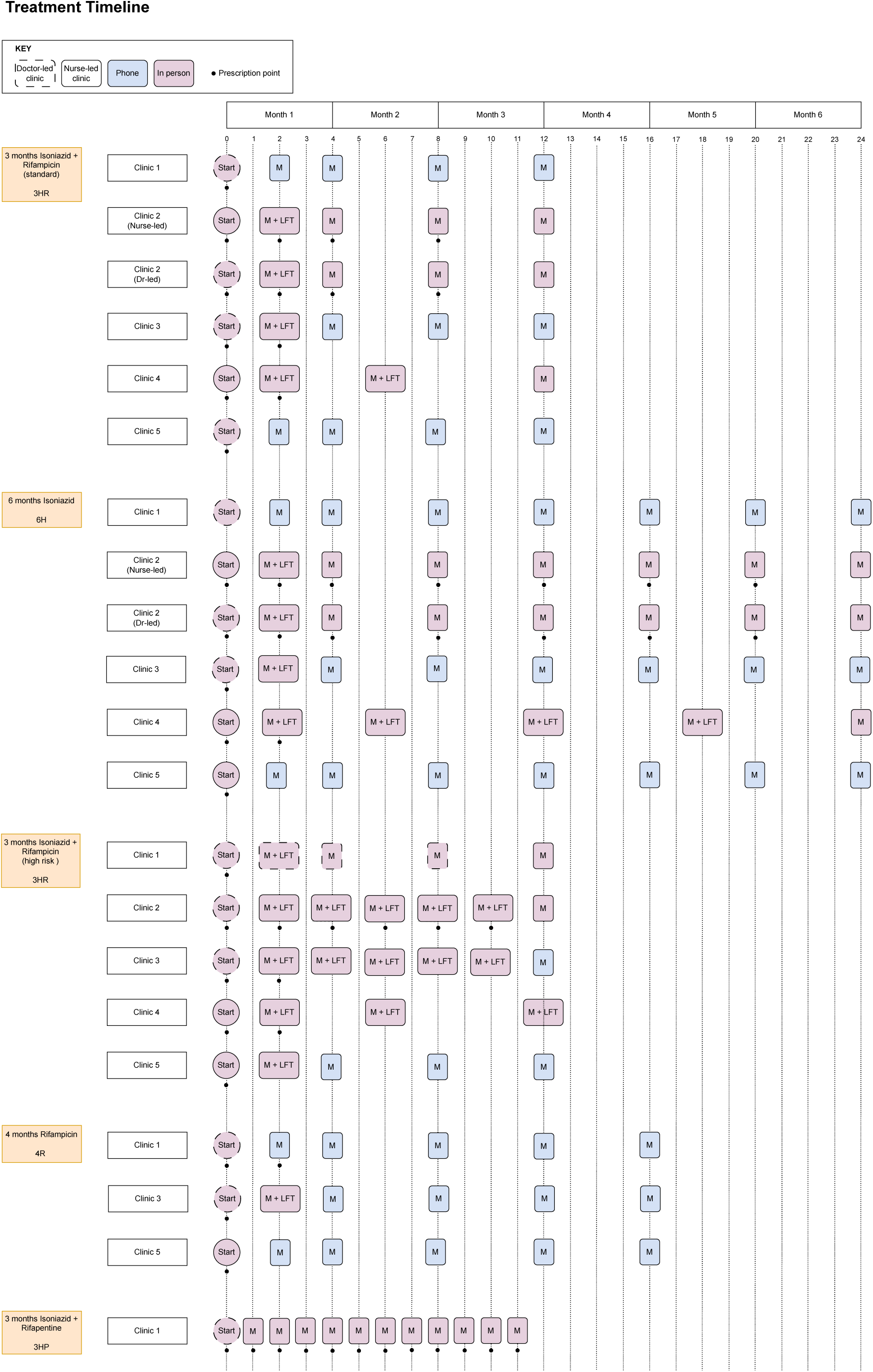
The weekly LTBI treatment and monitoring timeline for all five clinics and all regimens prescribed at each clinic.

For 3HR the mean cost of treatment was £192.68 (range: £140.43-£255.32), using UK salary costs. The two cheapest treatment pathways were nurse-led (Clinics 2 & 4), with the cheapest, Clinic 4, monitoring patients every six weeks rather than monthly. Clinic 4 is the only clinic to provide repeat monitoring tests, doing liver and renal function tests and complete blood count at every monitoring appointment except the final one; however, Clinic 4 reported consistently shorter appointments than the others. Clinics 1 & 5 do not routinely perform LFTs two weeks after treatment commencement.

For 6H and 4R the mean costs were £411.04(£362.56-£474.25) and £226.97(£190.15-£270.73), respectively. Here the monitoring processes followed the same pattern as 3HR but spanned six and four months respectively. For 6H, the drug costs (£259.10 compared to £78.88 for 3HR) considerably increased total cost.

For patients deemed high risk (3HR high-risk) the mean cost was £238.62 (range: £157.47-£346.03). Clinics 2 & 3 reported fortnightly monitoring with LFTs for patients deemed to be at higher risk of DILI, but said that in practice this could be as frequent as weekly depending on the level of risk. Clinic 1 conducts in-person monitoring with a consultant for high-risk patients, as opposed to nurse-led telephone monitoring as standard, while Clinic 5 retains telephone-based monitoring for high-risk patients but with an additional LFT at week 2. Clinic 4 conducts the same pattern of six-weekly liver and renal function tests and a full blood count for both standard and high-risk patients, but additionally repeated all tests on completion for the latter.

Three months of weekly Isoniazid and Rifapentine (3HP) is prescribed by Clinic 1 for patients with adherence concerns, at a cost of £559.05. These patients receive weekly in-person monitoring, which increases the staff time required, as well as the regimen’s being the most expensive, at an estimated cost of £300.

If a patient is lost to follow-up then costs are incurred without benefit, and the further the patient progresses along the pathway the greater the cost incurred. Figure 4 shows the weekly cumulative cost of 3HR, 3HR (high-risk), 6H and 4R regimens, taking into consideration when drugs are dispensed, staffing cost at monitoring appointments and the unit costs of any diagnostic tests done. As LTBI treatment costs are dominated by drug costs, clinics dispensing all drugs at week 0 (Clinics 1 & 5) or split between weeks 0 & 2 (Clinics 3 & 4) frontload their costs, while Clinic 2, which dispenses over time exhibits a steadier increase.

**Figure 4:**
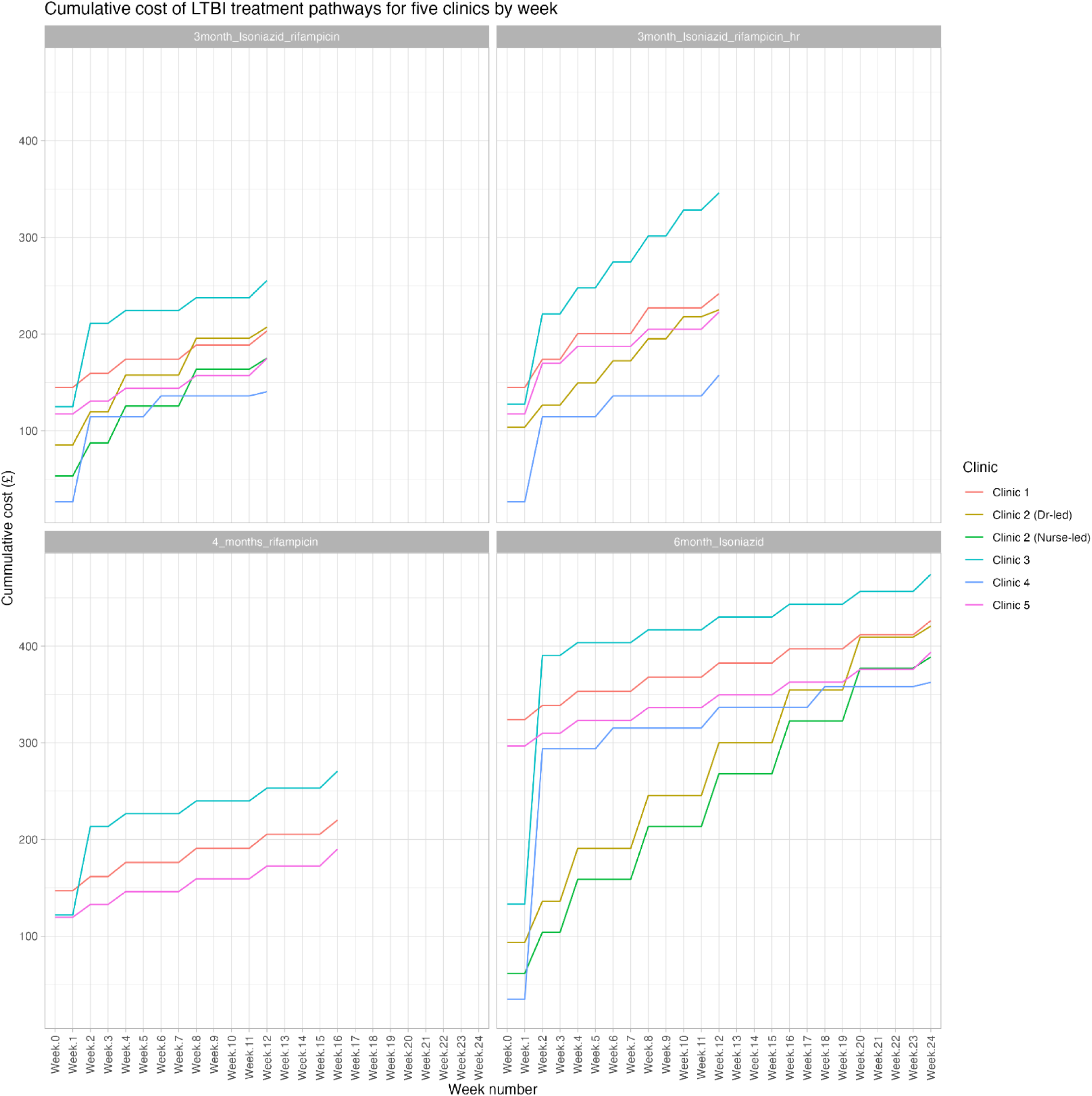
The cumulative cost of LTBI treatment at four clinics for 3HR, 3HR (higher risk patients), 4Rand 6H regimes by week. 2 has two routes: a nurse-led clinic (standard) and doctor-led (for patients with symptoms or deemed higher risk).

## DISCUSSION

To our knowledge this is the first detailed costing study of LTBI testing and treatment in the UK. We selected clinics with a geographic spread by sampling from the four TB Control Boards across England with the highest number of active TB cases, including two in London, which has the greatest number of TB cases in England.

Pathways differ amongst clinics for a variety of reasons, including the local configuration of services, and the particular needs of the patients they serve, and therefore variation in costs is to be expected. However, variation in costs associated with differences in clinical practice were modest, standard deviations being ≤22% of the mean for testing, and ≤29% for treatment.

Note that we calculated the costs for patients who complete the testing and treatment pathways. If patients are lost to follow-up, then some costs are incurred but for no benefit. Therefore, it is important that cost-effectiveness analyses using the information in this paper account for loss to follow-up. Public Health England reported in 2021 that “45% (921/2035) of people with a positive LTBI test were known to have accessed treatment in 2019 to 2020” and “where treatment data was available, during 2019 to 2020, 75% (480/637) of people were known to have completed treatment”^18^.

The analysis in this paper focuses on the patients who were screened and treated for LTBI (i.e. adult contacts of pulmonary TB cases and adult new entrants). Other groups are also recommended for screening and treatment, such as patients undergoing immunosuppressant treatment, healthcare workers with occupational exposure, children, and patients with comorbidities, and assessment of the cost-effectiveness of LTBI screening and treatment for those patients requires further work.

Additionally, primary care has an increasing role in LTBI testing and treatment, and analysis of those pathways in terms of costs and effectiveness is needed, as it may be cheaper and more accessible for some patients^19^.

## Data Availability

All data produced in the present study are available upon reasonable request to the authors

## Acknowledgment

We thank the Study Steering Committee for advice and support.

## FOOTNOTES

### Disclaimer

The views expressed are those of the authors and not necessarily those of the UK Department of Health and Social Care; Foreign, Commonwealth and Development Office; European Union; UK Medical Research Council (MRC); National Institute for Health and Care Research (NIHR); or UK Health Security Agency (UKHSA).

### Declaration of interests

PJW has received payment from Pfizer for teaching of mathematical modeling of infectious disease transmission and vaccination, and from the Dutch National Institute for Public Health and the Environment (RIVM) for participation in an audit committee on COVID-19 data analytics and modeling. All other authors report no potential conflicts.

**Table A1:** Costs of LTBI testing at four NHS TB clinics in England split by route through the pathway, using UK (non-London) payscales. Costs have been calculated for staff time, tests, and both combined. Average costs are unweighted means of the 4 clinics’ costs. See Appendix for calculations using Inner and Outer London payscales.

| Route | Staff Costs | Test costs | Total costs |
| --- | --- | --- | --- |
| **Asymptomatic IGRA-** |  |  |  |
| Clinic 1 | £43.61 | £26.00 | £69.61 |
| Clinic 2 | £31.73 | £25.00 | £56.73 |
| Clinic 3 | £51.02 | £30.00 | £81.02 |
| Clinic 4 | £24.08 | £25.00 | £49.08 |
| **Average** | **£37.61** | **£26.50** | **£64.11** |
| **Standard deviation** | **£12.02** | **£2.38** | **£14.10** |
| **Asymptomatic IGRA+** |  |  |  |
| Clinic 1 | £79.23 | £75.00 | £154.23 |
| Clinic 2 | £47.59 | £74.00 | £121.59 |
| Clinic 3 | £98.18 | £76.50 | £174.68 |
| Clinic 4 | £51.95 | £69.00 | £120.95 |
| **Average** | **£69.24** | **£73.63** | **£142.86** |
| **Standard deviation** | **£23.84** | **£3.25** | **£26.29** |
| **Symptomatic IGRA-** |  |  |  |
| Clinic 1 | £56.85 | £75.00 | £131.85 |
| Clinic 2 | £52.01 | £74.00 | £126.01 |
| Clinic 3 | £61.73 | £76.50 | £138.23 |
| Clinic 4 | £32.29 | £69.00 | £101.29 |
| **Average** | **£50.72** | **£73.63** | **£124.35** |
| **Standard deviation** | **£12.91** | **£3.25** | **£16.16** |
| **Symptomatic IGRA+** |  |  |  |
| Clinic 1 | £56.85 | £75.00 | £131.85 |
| Clinic 2 | £52.01 | £74.00 | £126.01 |
| Clinic 3 | £61.73 | £76.50 | £138.23 |
| Clinic 4 | £32.29 | £69.00 | £101.29 |
| **Average** | **£50.72** | **£73.63** | **£124.35** |
| **Standard deviation** | **£12.91** | **£3.25** | **£16.16** |

**Table A2:**
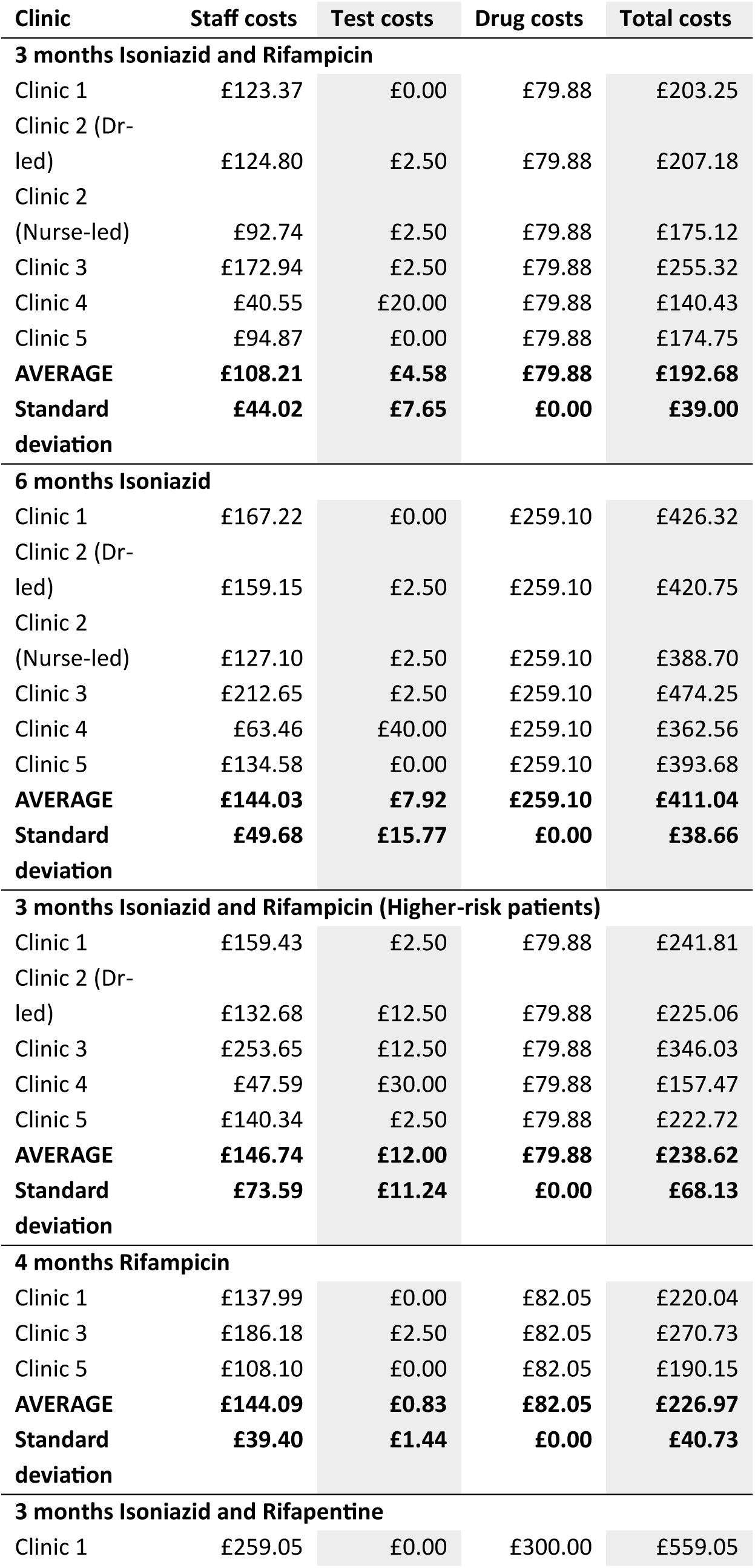
Costs of LTBI Treatment at five NHS TB clinics in England split by regimen, using UK (non-London) payscales. Costs have been calculated for staff time, tests, drugs, and all three combined. Average costs are unweighted means of the 5 clinics’ costs. Some clinics do not perform testing during treatment, so their test costs are zero. See Appendix for calculations using Inner and Outer London payscales.

## APPENDIX

### 1. Staff time – cost per minute

The cost per minute of staff time was calculated as described below.

#### Salary

Salary information 2023/2024was collected from the Nursing Pay Guide from Nurses.co.uk^1^ and from either Health Careers on the NHS Careers Website^2^ or the Pay and Contracts section of the BMA^3,4^. The midpoint salary for each grade was used. Payscales differ by geographic location, with three scales: for the UK except London, Outer London, and Inner London^5,6^.

#### Employer’s pension contribution

The employer pension contributions have been calculated at 23.78% (inclusive of the administration charge), which is the rate from 2023-2024^7^.

#### Employer’s National Insurance (NI) contribution

National Insurance contributions have been calculated using information from Gov.uk. For 2023-24 these contributions are 13.8% of an employee’s salary beyond the threshold of £9,100^8^.

#### Apprenticeship Levy

The Apprenticeship Levy is a tax paid by employers with total annual salary costs exceeding £3 million, and is charged at 0.5% of the employer’s salary costs^9^. Therefore, the per-person cost is 0.5% of salary.

#### Indirect costs

Indirect costs include overheads such as heating, lighting, building maintenance, auxiliary staffing requirements for administration, IT, humans resources and management. NIHR costing guidance^10^ estimated this to be 70% of NHS staff direct costs, so this was applied to Salary + Employer’s pension contribution + Employer’s NI contribution + Apprenticeship Levy.

#### Minutes worked per year

There are 260 weekdays in a year, and 8 public holidays. Days worked per year for nursing staff and foundation level doctors have been calculated assuming a holiday entitlement of 27 days + 8 public holidays, meaning they work 225 days a year. Days worked per year for doctors at CMT, Registrar, Consultant and Associate Specialist have been calculated assuming a 30-day holiday entitlement + 8 public holidays, meaning they work 222 days^11^. Nurses’ salaries are calculated on a working day of 7.5 hours (37.5 hours per week); for doctors there is a 40-hour week (5 × 8 hours). The total employment cost was then divided by minutes worked per year to give a cost per minute for each staff grade.

**Table A3:** Per-minute staffing cost calculations using salary information for the UK (excluding London).

| Type of staff | Salary | Pension<br>(employer's<br>contribution) | NI<br>(employer's<br>contribution) | Apprenticeship<br>levy | Indirect costs<br>(70% of<br>employment<br>costs) | Total<br>employment<br>cost | Minutes worked<br>per year | Cost per<br>minute |
| --- | --- | --- | --- | --- | --- | --- | --- | --- |
| **Doctors** |  |  |  |  |  |  |  |  |
| Foundation | £34,851 | £8,287 | £3,554 | £174 | £32,806 | £79,672 | 108,000 | £0.74 |
| CMT | £49,626 | £11,801 | £5,593 | £248 | £47,087 | £114,355 | 106,560 | £1.07 |
| Registrar | £53,538 | £12,731 | £6,132 | £268 | £50,868 | £123,537 | 106,560 | £1.16 |
| Consultant | £109,974 | £26,152 | £13,921 | £550 | £105,417 | £256,013 | 106,560 | £2.40 |
| Associate Specialist | £87,106 | £20,714 | £10,765 | £436 | £83,314 | £202,334 | 106,560 | £1.90 |
| **Nurses** |  |  |  |  |  |  |  |  |
| Band 3 | £23,576 | £5,606 | £1,998 | £118 | £21,909 | £53,206 | 101,250 | £0.53 |
| Band 4 | £26,372 | £6,271 | £2,383 | £132 | £24,611 | £59,769 | 101,250 | £0.59 |
| Band 5 | £34,581 | £8,223 | £3,516 | £173 | £32,546 | £79,039 | 101,250 | £0.78 |
| Band 6 | £38,974 | £9,268 | £4,123 | £195 | £36,791 | £89,350 | 101,250 | £0.88 |
| Band 7 | £46,899 | £11,153 | £5,216 | £234 | £44,452 | £107,954 | 101,250 | £1.07 |
| Band 8a | £54,151 | £12,877 | £6,217 | £271 | £51,461 | £124,976 | 101,250 | £1.23 |
| **Other Healthcare Workers** |  |  |  |  |  |  |  |  |
| Admin Support Worker (Band 3) | £23,576 | £5,606 | £1,998 | £118 | £21,909 | £53,206 | 101,250 | £0.53 |
| Healthcare Support Worker (Band 3) | £23,576 | £5,606 | £1,998 | £118 | £21,909 | £53,206 | 101,250 | £0.53 |
| Phlebotomist (Band 3) | £23,576 | £5,606 | £1,998 | £118 | £21,909 | £53,206 | 101,250 | £0.53 |
| Pharmacist (Band 6) | £38,974 | £9,268 | £4,123 | £195 | £36,791 | £89,350 | 101,250 | £0.88 |
| Radiographer (Band 6) | £38,974 | £9,268 | £4,123 | £195 | £36,791 | £89,350 | 101,250 | £0.88 |

**Table A4:** Per-minute staffing cost calculations using salary information for Outer London.

| Type of staff | Salary | Outer London supplement | Total salary | Pension (employer's contribution) | NI (employer's contribution) | Apprenticeship levy | Indirect costs (70% of employment costs) | Total employment cost | Minutes worked per year | Cost per minute |
| --- | --- | --- | --- | --- | --- | --- | --- | --- | --- | --- |
| Doctors |  |  |  |  |  |  |  |  |  |  |
| Foundation | £34,851 | £527 | £35,378 | £8,413 | £3,626 | £177 | £33,316 | £80,910 | 108,000 | £0.75 |
| CMT | £49,626 | £527 | £50,153 | £11,926 | £5,665 | £251 | £47,597 | £115,592 | 106,560 | £1.08 |
| Registrar | £53,538 | £527 | £54,065 | £12,857 | £6,205 | £270 | £51,378 | £124,775 | 106,560 | £1.17 |
| Consultant | £109,974 | £527 | £110,501 | £26,277 | £13,993 | £553 | £105,927 | £257,251 | 106,560 | £2.41 |
| Associate Specialist | £87,106 | £527 | £87,633 | £20,839 | £10,838 | £438 | £83,823 | £203,571 | 106,560 | £1.91 |
| Nurses |  |  |  |  |  |  |  |  |  |  |
| Band 3 | £23,576 | £4,314 | £27,890 | £6,632 | £2,593 | £139 | £26,078 | £63,333 | 101,250 | £0.63 |
| Band 4 | £26,372 | £4,314 | £30,686 | £7,297 | £2,979 | £153 | £28,781 | £69,896 | 101,250 | £0.69 |
| Band 5 | £34,581 | £5,187 | £39,768 | £9,457 | £4,232 | £199 | £37,559 | £91,215 | 101,250 | £0.90 |
| Band 6 | £38,974 | £5,436 | £44,410 | £10,561 | £4,873 | £222 | £42,046 | £102,111 | 101,250 | £1.01 |
| Band 7 | £46,899 | £5,436 | £52,335 | £12,445 | £5,966 | £262 | £49,706 | £120,714 | 101,250 | £1.19 |
| Band 8 a | £54,151 | £5,436 | £59,587 | £14,170 | £6,967 | £298 | £56,715 | £137,737 | 101,250 | £1.36 |
| Other Healthcare Workers |  |  |  |  |  |  |  |  |  |  |
| Admin Support Worker (Band 3) | £23,576 | £4,314 | £27,890 | £6,632 | £2,593 | £139 | £26,078 | £63,333 | 101,250 | £0.63 |
| Healthcare Support Worker (Band 3) | £23,576 | £4,314 | £27,890 | £6,632 | £2,593 | £139 | £26,078 | £63,333 | 101,250 | £0.63 |
| Phlebotomist (Band 3) | £23,576 | £4,314 | £27,890 | £6,632 | £2,593 | £139 | £26,078 | £63,333 | 101,250 | £0.63 |
| Pharmacist (Band 6) | £38,974 | £5,436 | £44,410 | £10,561 | £4,873 | £222 | £42,046 | £102,111 | 101,250 | £1.01 |
| Radiographer (Band 6) | £38,974 | £5,436 | £44,410 | £10,561 | £4,873 | £222 | £42,046 | £102,111 | 101,250 | £1.01 |

**Table A5:** Per-minute staffing cost calculations using salary information for Inner London.

| Type of staff | Salary | Inner London supplement | Total salary | Pension (employer's contribution) | NI (employer's contribution) | Apprenticeship levy | Indirect costs (70% of employment costs) | Total employment cost | Minutes worked per year | Cost per minute |
| --- | --- | --- | --- | --- | --- | --- | --- | --- | --- | --- |
| Doctors |  |  |  |  |  |  |  |  |  |  |
| Foundation | £34,851 | £2,162 | £37,013 | £8,802 | £3,852 | £185 | £34,896 | £84,747 | 108,000 | £0.78 |
| CMT | £49,626 | £2,162 | £51,788 | £12,315 | £5,891 | £259 | £49,177 | £119,430 | 106,560 | £1.12 |
| Registrar | £53,538 | £2,162 | £55,700 | £13,245 | £6,431 | £278 | £52,958 | £128,612 | 106,560 | £1.21 |
| Consultant | £109,974 | £2,162 | £112,136 | £26,666 | £14,219 | £561 | £107,507 | £261,088 | 106,560 | £2.45 |
| Associate Specialist | £87,106 | £2,162 | £89,268 | £21,228 | £11,063 | £446 | £85,404 | £207,409 | 106,560 | £1.95 |
| Nurses |  |  |  |  |  |  |  |  |  |  |
| Band 3 | £23,576 | £5,132 | £28,708 | £6,827 | £2,706 | £144 | £26,869 | £65,253 | 101,250 | £0.64 |
| Band 4 | £26,372 | £5,274 | £31,646 | £7,525 | £3,111 | £158 | £29,709 | £72,149 | 101,250 | £0.71 |
| Band 5 | £34,581 | £6,916 | £41,497 | £9,868 | £4,471 | £207 | £39,230 | £95,274 | 101,250 | £0.94 |
| Band 6 | £38,974 | £7,746 | £46,720 | £11,110 | £5,191 | £234 | £44,278 | £107,533 | 101,250 | £1.06 |
| Band 7 | £46,899 | £7,746 | £54,645 | £12,995 | £6,285 | £273 | £51,939 | £126,137 | 101,250 | £1.25 |
| Band 8 a | £54,151 | £7,746 | £61,897 | £14,719 | £7,286 | £309 | £58,948 | £143,159 | 101,250 | £1.41 |
| Other Healthcare Workers |  |  |  |  |  |  |  |  |  |  |
| Admin Support Worker (Band 3) | £23,576 | £5,132 | £28,708 | £6,827 | £2,706 | £144 | £26,869 | £65,253 | 101,250 | £0.64 |
| Healthcare Support Worker (Band 3) | £23,576 | £5,132 | £28,708 | £6,827 | £2,706 | £144 | £26,869 | £65,253 | 101,250 | £0.64 |
| Phlebotomist (Band 3) | £23,576 | £5,132 | £28,708 | £6,827 | £2,706 | £144 | £26,869 | £65,253 | 101,250 | £0.64 |
| Pharmacist (Band 6) | £38,974 | £7,746 | £46,720 | £11,110 | £5,191 | £234 | £44,278 | £107,533 | 101,250 | £1.06 |
| Radiographer (Band 6) | £38,974 | £7,746 | £46,720 | £11,110 | £5,191 | £234 | £44,278 | £107,533 | 101,250 | £1.06 |

### 2. Drug regimen costs

Five drug regimens to treat LTBI are currently in use at the clinics sampled. Costs for each regimen have been calculated using costs data from the British National Formulary^12^or EMIT database^13^, selecting the cheapest option from either. First prescribing guidelines from NICE were compared with data from clinicians for each regimen to establish a daily dosage for each drug prescribed. As dosage varies dependent on weight, an adult of 70kg was chosen as an example case. Then using daily dosage data, regimen length, and information on pack size and cost, it was possible to calculate a total cost for each drugs regimen.

**Table A6:**
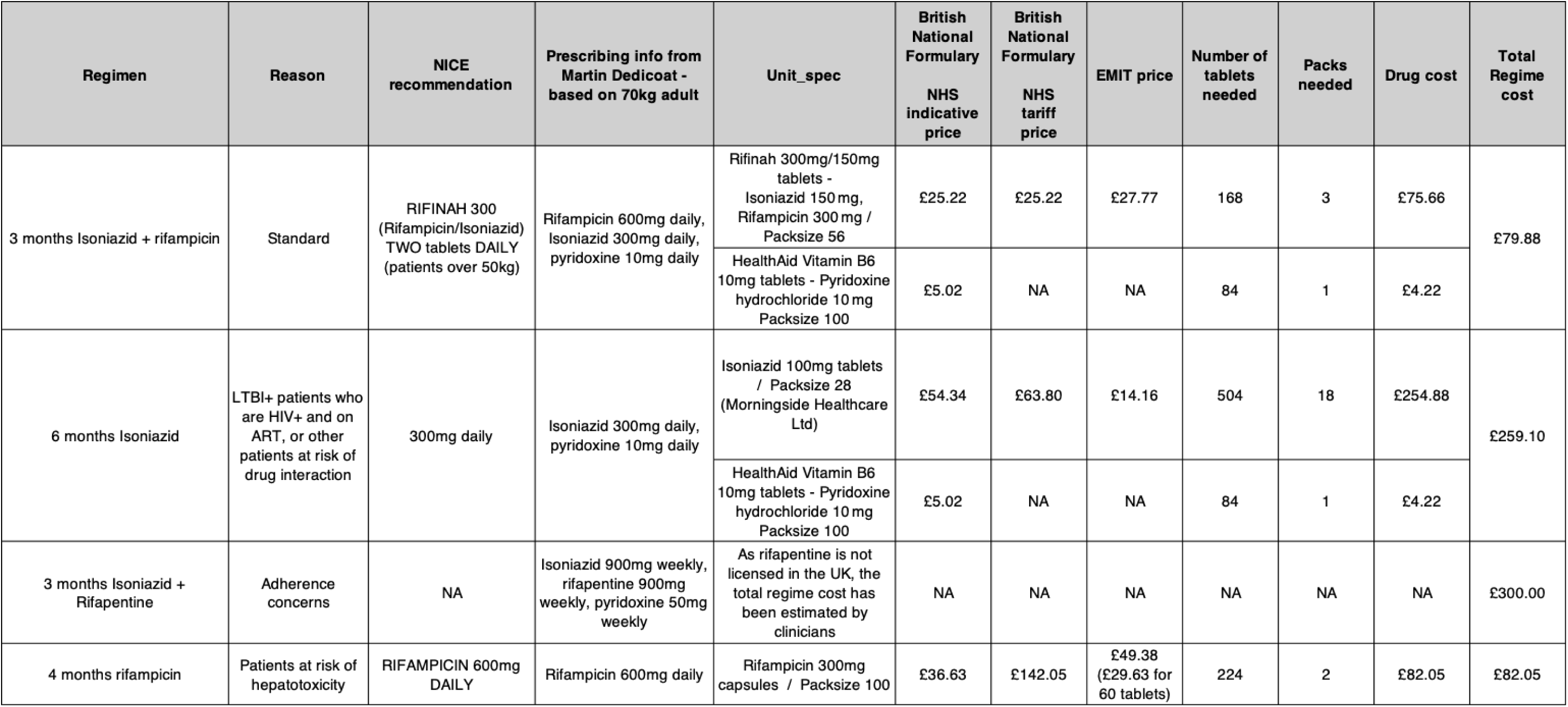
Drugs costs used in the LTBI treatment calculations.

### 3. Clinical pathways

The pathways analysed in this paper are below.

**Figure A1:**
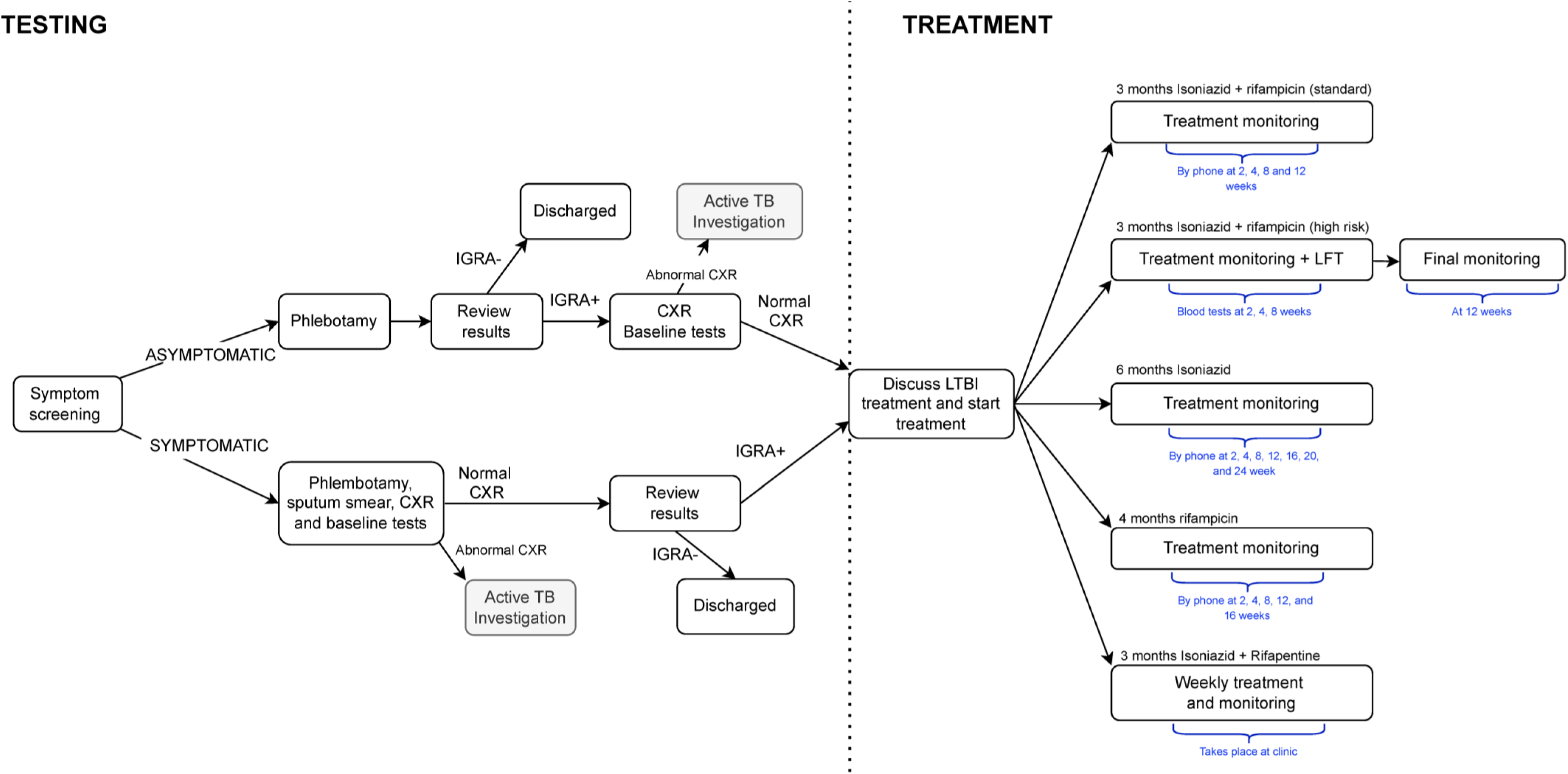
LTBI testing and treatment pathways at Clinic 1.

**Figure A2:**
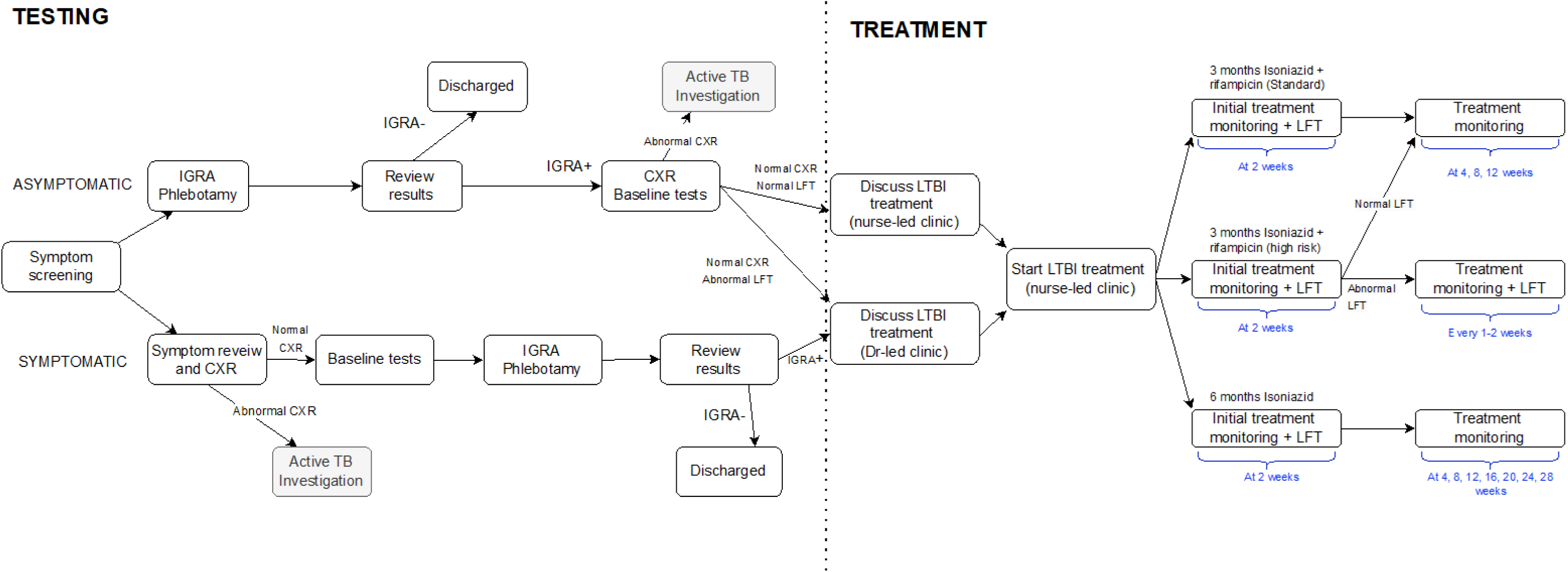
LTBI testing and treatment pathways at Clinic 2. A decision to consider the nurse-led and doctor-led treatment routes separately was made to capture the different cost implications of both routes.

**Figure A3:**
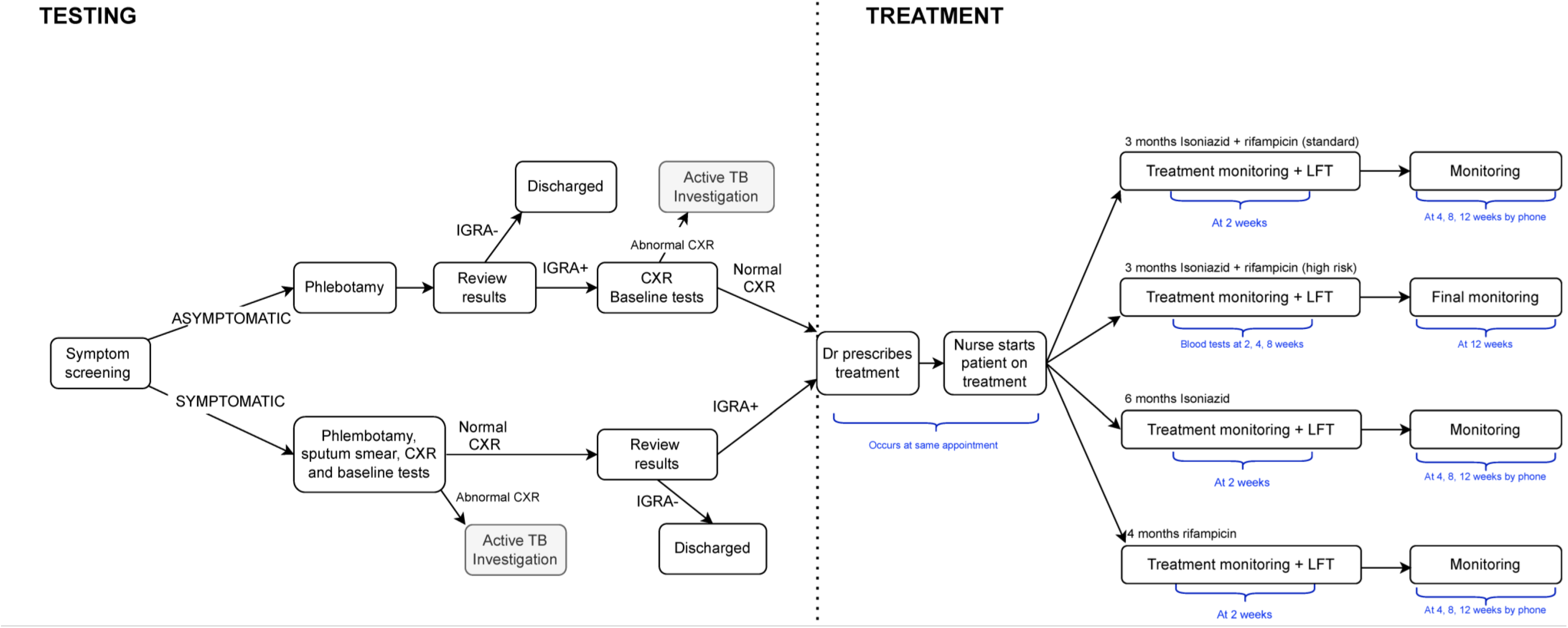
LTBI testing and treatment pathways at Clinic 3.

**Figure A4:**
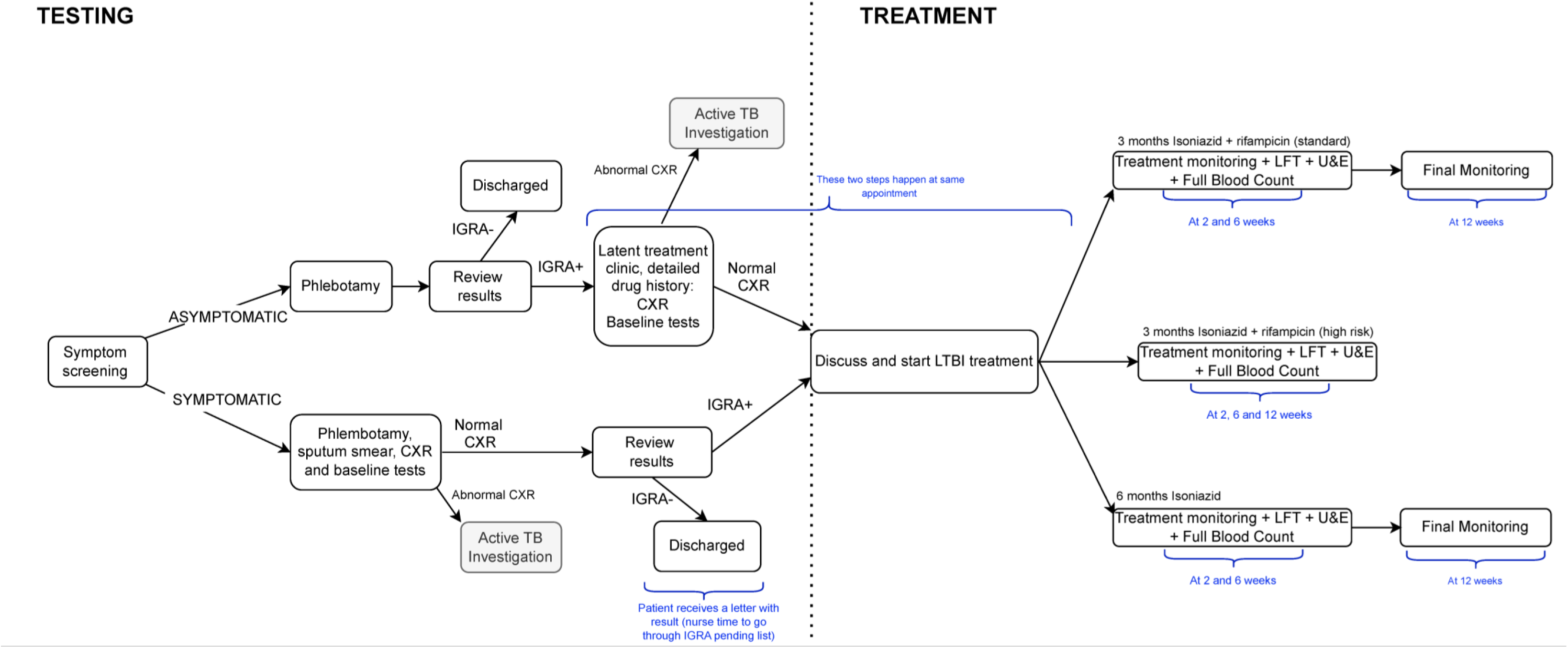
LTBI testing and treatment pathways at Clinic 4.

**Figure A5:**
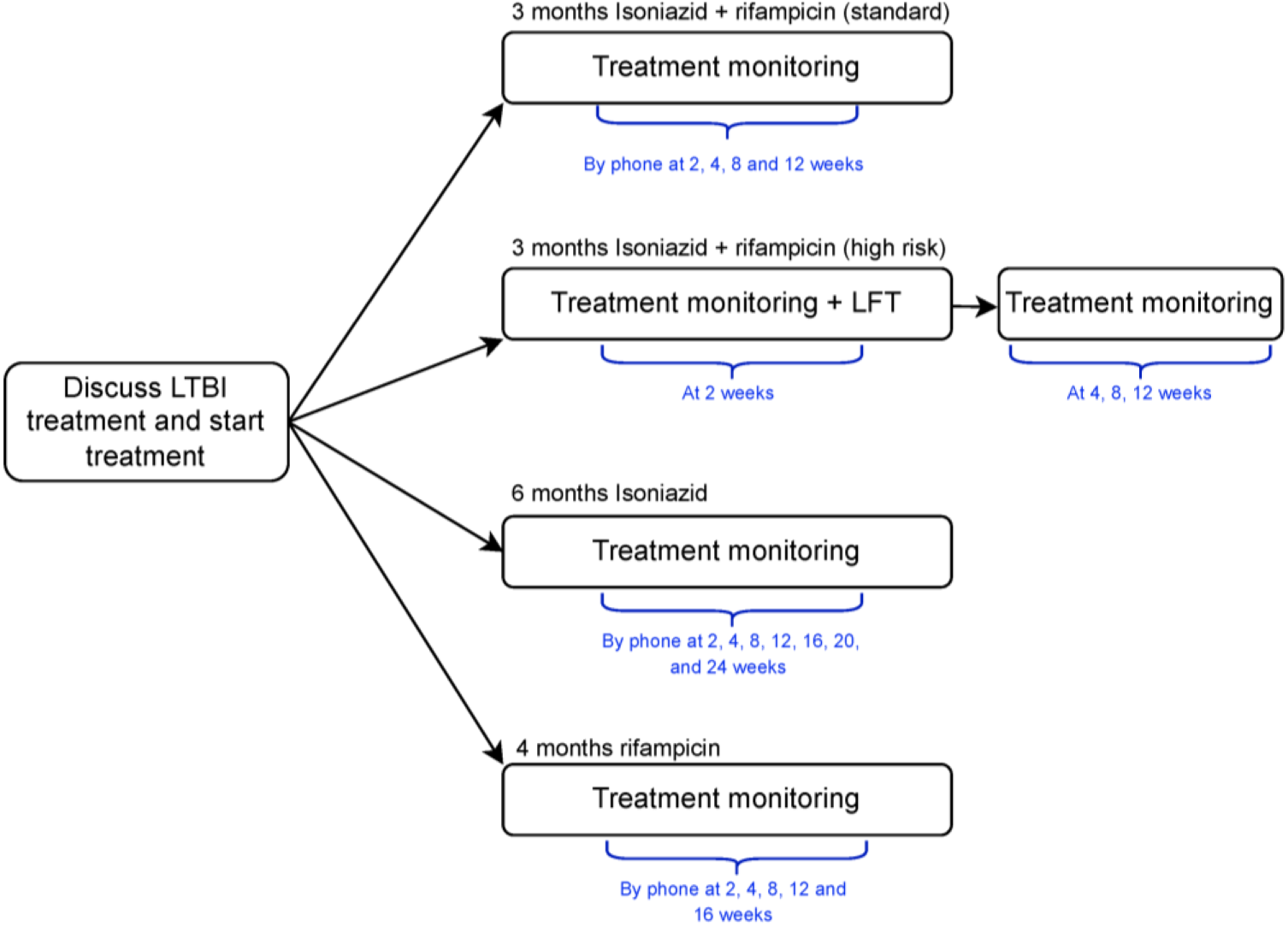
LTBI treatment pathway at Clinic 5. The testing pathways includes primary care and was not considered in this analysis.

### 4. Testing and treatment costs for Inner and Outer London

**Table A5:** Costs of LTBI testing at four NHS TB clinics in England split by route through the pathway, using Outer London pay scales. Costs have been calculated for staff time, tests, and both combined. Average costs are unweighted means of the 4 clinics’ costs.

| Route | Staff Costs | Test costs | Total costs |
| --- | --- | --- | --- |
| **Asymptomatic IGRA-** |  |  |  |
| Clinic 1 | £47.28 | £26.00 | £73.28 |
| Clinic 2 | £36.51 | £25.00 | £61.51 |
| Clinic 3 | £59.54 | £30.00 | £89.54 |
| Clinic 4 | £26.81 | £25.00 | £51.81 |
| **Average** | **£42.54** | **£26.50** | **£69.04** |
| **Standard deviation** | **£14.09** | **£2.38** | **£16.25** |
| **Asymptomatic IGRA+** |  |  |  |
| Clinic 1 | £83.13 | £75.00 | £158.13 |
| Clinic 2 | £54.77 | £74.00 | £128.77 |
| Clinic 3 | £108.92 | £76.50 | £185.42 |
| Clinic 4 | £57.14 | £69.00 | £126.14 |
| **Average** | **£75.99** | **£73.63** | **£149.62** |
| **Standard deviation** | **£25.44** | **£3.25** | **£27.93** |
| **Symptomatic IGRA-** |  |  |  |
| Clinic 1 | £62.41 | £75.00 | £137.41 |
| Clinic 2 | £59.81 | £74.00 | £133.81 |
| Clinic 3 | £71.03 | £76.50 | £147.53 |
| Clinic 4 | £35.37 | £69.00 | £104.37 |
| **Average** | **£57.16** | **£73.63** | **£130.78** |
| **Standard deviation** | **£15.29** | **£3.25** | **£18.54** |
| **Symptomatic IGRA+** |  |  |  |
| Clinic 1 | £62.41 | £75.00 | £137.41 |
| Clinic 2 | £59.81 | £74.00 | £133.81 |
| Clinic 3 | £71.03 | £76.50 | £147.53 |
| Clinic 4 | £35.37 | £69.00 | £104.37 |
| **Average** | **£57.16** | **£73.63** | **£130.78** |
| **Standard deviation** | **£15.29** | **£3.25** | **£18.54** |

**Table A6:** Costs of LTBI testing at four NHS TB clinics in England split by route through the pathway, using Inner London pay scales. Costs have been calculated for staff time, tests, and both combined. Average costs are unweighted means of the 4 clinics’ costs.

| Route | Staff Costs | Test costs | Total costs |
| --- | --- | --- | --- |
| **Asymptomatic IGRA-** |  |  |  |
| Clinic 1 | £48.99 | £26.00 | £74.99 |
| Clinic 2 | £38.31 | £25.00 | £63.31 |
| Clinic 3 | £61.75 | £30.00 | £91.75 |
| Clinic 4 | £27.93 | £25.00 | £52.93 |
| **Average** | **£44.25** | **£26.50** | **£70.75** |
| **Standard deviation** | **£14.50** | **£2.38** | **£16.65** |
| **Asymptomatic IGRA+** |  |  |  |
| Clinic 1 | £85.56 | £75.00 | £160.56 |
| Clinic 2 | £57.46 | £74.00 | £131.46 |
| Clinic 3 | £113.24 | £76.50 | £189.74 |
| Clinic 4 | £59.34 | £69.00 | £128.34 |
| **Average** | **£78.90** | **£73.63** | **£152.53** |
| **Standard deviation** | **£26.24** | **£3.25** | **£28.74** |
| **Symptomatic IGRA-** |  |  |  |
| Clinic 1 | £64.92 | £75.00 | £139.92 |
| Clinic 2 | £62.77 | £74.00 | £136.77 |
| Clinic 3 | £74.27 | £76.50 | £150.77 |
| Clinic 4 | £36.71 | £69.00 | £105.71 |
| **Average** | **£59.67** | **£73.63** | **£133.29** |
| **Standard deviation** | **£16.10** | **£3.25** | **£19.34** |
| **Symptomatic IGRA+** |  |  |  |
| Clinic 1 | £64.92 | £75.00 | £139.92 |
| Clinic 2 | £62.77 | £74.00 | £136.77 |
| Clinic 3 | £74.27 | £76.50 | £150.77 |
| Clinic 4 | £36.71 | £69.00 | £105.71 |
| **Average** | **£59.67** | **£73.63** | **£133.29** |
| **Standard deviation** | **£16.10** | **£3.25** | **£19.34** |

**Table A7:** Costs of LTBI treatment at five NHS TB clinics in England split by regimen, using Outer London pay scales. Costs have been calculated for staff time, tests, drugs, and all three combined. Average costs are unweighted means of the 5 clinics’ costs. Some clinics do not perform testing during treatment, so their test costs are zero.

| Clinic | Staff costs | Test costs | Drug costs | Total costs |
| --- | --- | --- | --- | --- |
| **3 months Isoniazid and Rifampicin** |  |  |  |  |
| Clinic 1 | £136.18 | £0.00 | £79.88 | £216.06 |
| Clinic 2 (Dr-led) | £136.35 | £2.50 | £79.88 | £218.73 |
| Clinic 2 (Nurse-led) | £105.95 | £2.50 | £79.88 | £188.33 |
| Clinic 3 | £192.69 | £2.50 | £79.88 | £275.07 |
| Clinic 4 | £46.60 | £20.00 | £79.88 | £146.48 |
| Clinic 5 | £108.41 | £0.00 | £79.88 | £188.29 |
| **AVERAGE** | **£121.03** | **£4.58** | **£79.88** | **£205.49** |
| **Standard deviation** | **£48.01** | **£7.65** | **£0.00** | **£42.90** |
| **6 months Isoniazid** |  |  |  |  |
| Clinic 1 | £185.69 | £0.00 | £259.10 | £444.79 |
| Clinic 2 (Dr-led) | £175.99 | £2.50 | £259.10 | £437.59 |
| Clinic 2 (Nurse-led) | £145.59 | £2.50 | £259.10 | £407.19 |
| Clinic 3 | £238.07 | £2.50 | £259.10 | £499.67 |
| Clinic 4 | £73.02 | £40.00 | £259.10 | £372.12 |
| Clinic 5 | £153.80 | £0.00 | £259.10 | £412.90 |
| **AVERAGE** | **£162.03** | **£7.92** | **£259.10** | **£429.04** |
| **Standard deviation** | **£54.40** | **£15.77** | **£0.00** | **£43.10** |
| **3 months Isoniazid and Rifampicin (Higher-risk patients)** |  |  |  |  |
| Clinic 1 | £170.69 | £2.50 | £79.88 | £253.07 |
| Clinic 2 (Dr-led) | £145.74 | £12.50 | £79.88 | £238.12 |
| Clinic 3 | £283.48 | £12.50 | £79.88 | £375.86 |
| Clinic 4 | £54.77 | £30.00 | £79.88 | £164.65 |
| Clinic 5 | £156.24 | £2.50 | £79.88 | £238.62 |
| **AVERAGE** | **£162.18** | **£12.00** | **£79.88** | **£254.06** |
| **Standard deviation** | **£81.59** | **£11.24** | **£0.00** | **£76.36** |
| **4 months Rifampicin** |  |  |  |  |
| Clinic 1 | £152.68 | £0.00 | £82.05 | £234.73 |
| Clinic 3 | £207.82 | £2.50 | £82.05 | £292.37 |
| Clinic 5 | £123.54 | £0.00 | £82.05 | £205.59 |
| **AVERAGE** | **£161.35** | **£0.83** | **£82.05** | **£244.23** |
| **Standard deviation** | **£42.80** | **£1.44** | **£0.00** | **£44.16** |
| **3 months Isoniazid and Rifapentine** |  |  |  |  |
| Clinic 1 | £292.03 | £0.00 | £300.00 | £592.03 |

**Table A8:**
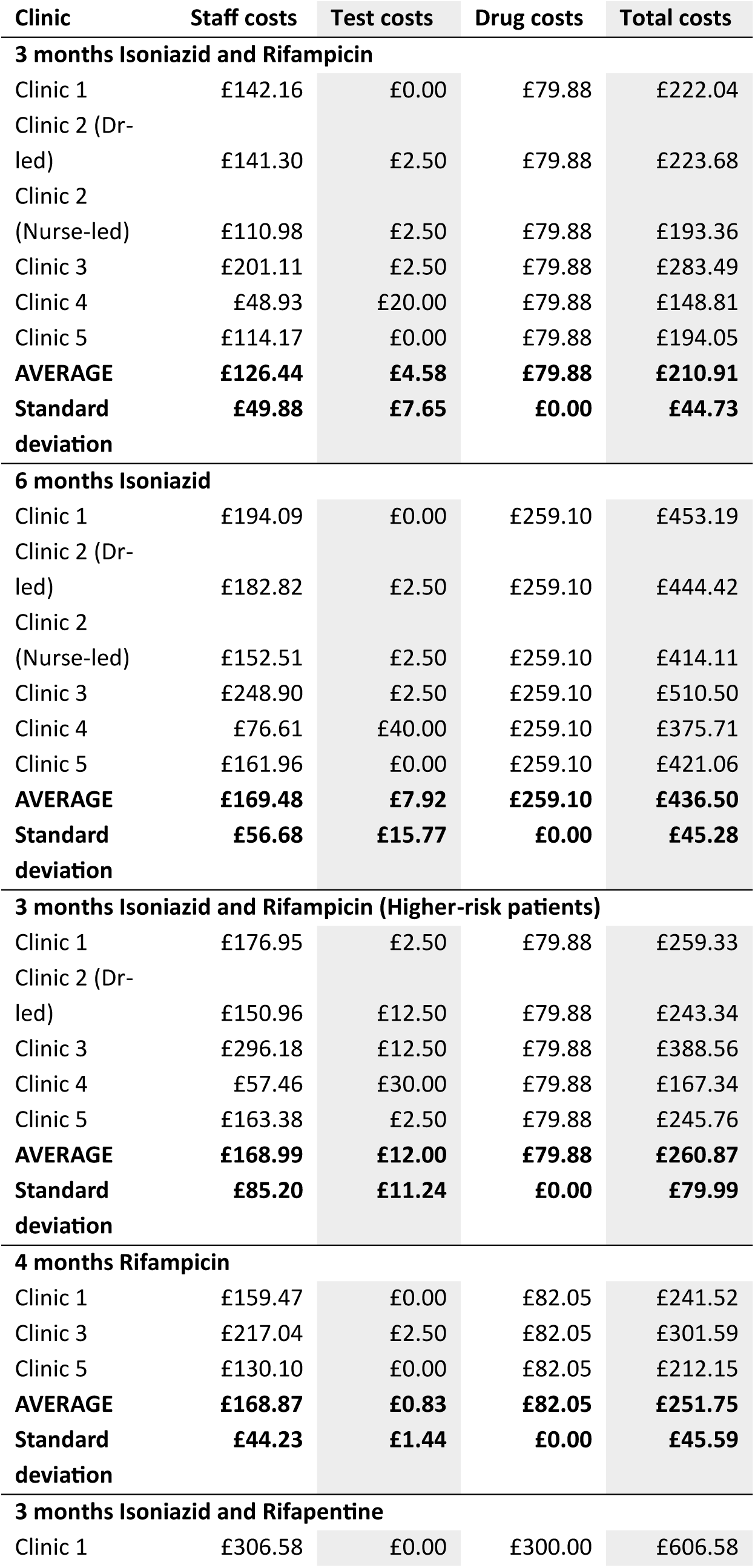
Costs of LTBI treatment at five NHS TB clinics in England split by regimen, using Inner London pay scales. Costs have been calculated for staff time, tests, drugs, and all three combined. Average costs are unweighted means of the 5 clinics’ costs. Some clinics do not perform testing during treatment, so their test costs are zero.

